# Deep learning based electrocardiographic screening for chronic kidney disease

**DOI:** 10.1101/2022.03.01.22271473

**Authors:** Lauri Holmstrom, Matthew Christensen, Neal Yuan, J. Weston Hughes, John Theurer, Melvin Jujjavarapu, Pedram Fatehi, Alan Kwan, Roopinder K. Sandhu, Joseph Ebinger, Susan Cheng, James Zou, Sumeet S. Chugh, David Ouyang

## Abstract

**Background:** Undiagnosed chronic kidney disease (CKD) is a common and usually asymptomatic disorder that causes a high burden of morbidity and early mortality worldwide. We developed a deep learning model for CKD screening from routinely acquired ECGs.

**Methods:** We collected data from a primary cohort with 111,370 patients which had 247,655 ECGs between 2005 and 2019. Using this data, we developed, trained, validated, and tested a deep learning model to predict whether an ECG was taken within one year of the patient receiving a CKD diagnosis. The model was additionally validated using an external cohort from another healthcare system which had 312,145 patients with 896,620 ECGs from between 2005 and 2018.

**Results:** Using 12-lead ECG waveforms, our deep learning algorithm achieved discrimination for CKD of any stage with an AUC of 0.77 (95% CI 0.76-0.77) in a held-out test set and an AUC of 0.71 (0.71-0.71) in the external cohort. Our 12-lead ECG-based model performance was consistent across the severity of CKD, with an AUC of 0.75 (0.0.74-0.77) for mild CKD, AUC of 0.76 (0.75-0.77) for moderate-severe CKD, and an AUC of 0.78 (0.77-0.79) for ESRD. In our internal health system with 1-lead ECG waveform data, our model achieved an AUC of 0.74 (0.74-0.75) in detecting any stage CKD. In the external cohort, our 1-lead ECG-based model achieved an AUC of 0.70 (0.70-0.70). In patients under 60 years old, our model achieved high performance in detecting any stage CKD with both 12-lead (AUC 0.84 [0.84-0.85]) and 1-lead ECG waveform (0.82 [0.81-0.83]).

**Conclusions:** Our deep learning algorithm was able to detect CKD using ECG waveforms, with particularly strong performance in younger patients and patients with more severe stages of CKD. Given the high global burden of undiagnosed CKD, further studies are warranted to evaluate the clinical utility of ECG-based CKD screening.

## Introduction

Almost 700 million individuals globally have chronic kidney disease (CKD), an important but often unrecognized cause of morbidity and early mortality (1). The initial presentation of CKD is usually asymptomatic and without overt clinical manifestations especially in early stages of the disease. Recently, the Global Burden of Diseases, Injuries and Risk Factors Study (GBD) estimated that CKD accounts for 4.6% of total mortality worldwide, with a 41.5% increase between 1990-2017(1). Delayed diagnosis and limited patient recognition of the condition contributes significantly to the burden of morbidity(2, 3). Early detection can potentially change the disease trajectory. The most common causes of CKD, such as hypertension and diabetes, can be reversible or treatable, and early diagnosis is crucial for avoiding renal replacement therapy(4, 5). There are few methods to cheaply or non-invasively screen for CKD, with conventional risk calculators lacking specificity and requiring both serum and urine laboratory testing(6).

Electrocardiograms (ECGs) are inexpensive, non-invasive, widely available, and rapid diagnostic tests frequently obtained during routine visits, prior to exercise, during preoperative evaluation, and for patients at increased risk of cardiovascular disease. Deep learning algorithms (DLA) have recently been applied to medical imaging and clinical data to achieve high precision, and to identify additional information beyond the interpretation of human experts(7, 8). Deep learning analysis of ECG waveforms has had potentially promising performance in prognosticating outcomes(9), identifying subclinical disease(10, 11), and identifying systemic phenotypes not traditionally associated with ECGs(12, 13). Given the prior success in identifying occult arrhythmias(14, 15), ventricular dysfunction(10), anemia(13), and age(12), DLA applied to screening ECGs could potentially identify patients who would benefit from further evaluation for kidney disease.

The high prevalence of concomitant cardiovascular disease and the well established changes that accompany electrolyte abnormalities suggest that the ECG is also altered in the setting of CKD and that discrete electrocardiographic signatures could be identifiable with deep learning techniques. Patients with CKD have a disproportionate accumulation of CV risk factors, such as diabetes and hypertension, as well as subclinical cardiovascular changes such as left ventricular hypertrophy, myocardial fibrosis, and diastolic dysfunction(16). It is not fully clear at which stage CKD patients start to develop manifest cardiovascular changes. However, recent studies have reported that patients with early-stage CKD may already have an increase in diffuse myocardial fibrosis on cardiac MRI(17). In addition to myocardial remodeling, CKD associates with a variety of electrolyte abnormalities that also cause widespread ECG abnormalities (e.g., decreased T-wave amplitudes in hypokalemia, large-amplitude T-waves, and prolonged QRS duration in hyperkalemia, and QTc prolongation in hypocalcemia)(18). Given such observations, it may be possible that asymptomatic CKD presents with subtle ECG alterations that are not visible to the human eye.

To overcome current limitations in screening for occult CKD, we designed, trained, and validated a deep learning model to predict CKD, including end-stage renal disease (ESRD), by analysis of waveform signals from a single 12-lead and 1-lead ECG. Incorporating both structured information from medical diagnoses as well as laboratory data, we assessed the ability of our model to evaluate the entire spectrum of kidney disease. To further evaluate our model, we validated its performance using corresponding data from a separate healthcare system.

## Methods

### Data sources and study population

We retrospectively identified 54,582 ECGs among 7,947 patients between 2005-2019 which were linked to a diagnosis of CKD within a 1-year window at Cedars-Sinai Medical Center. We also identified 193,073 ECGs among 103,814 patients between 2008-2019 with no CKD diagnoses at any point, which were used as matched negative controls. The study population from CSMC was randomly split 8:1:1 into training, validation, and test cohorts by patient such that the multiple ECGs from the same patient were limited to one cohort. In addition, we identified 896,620 ECGs among 312,145 patients at Stanford Healthcare from 8/2005 to 6/2018, which were used for external validation (Figure 1).

**Figure 1.**
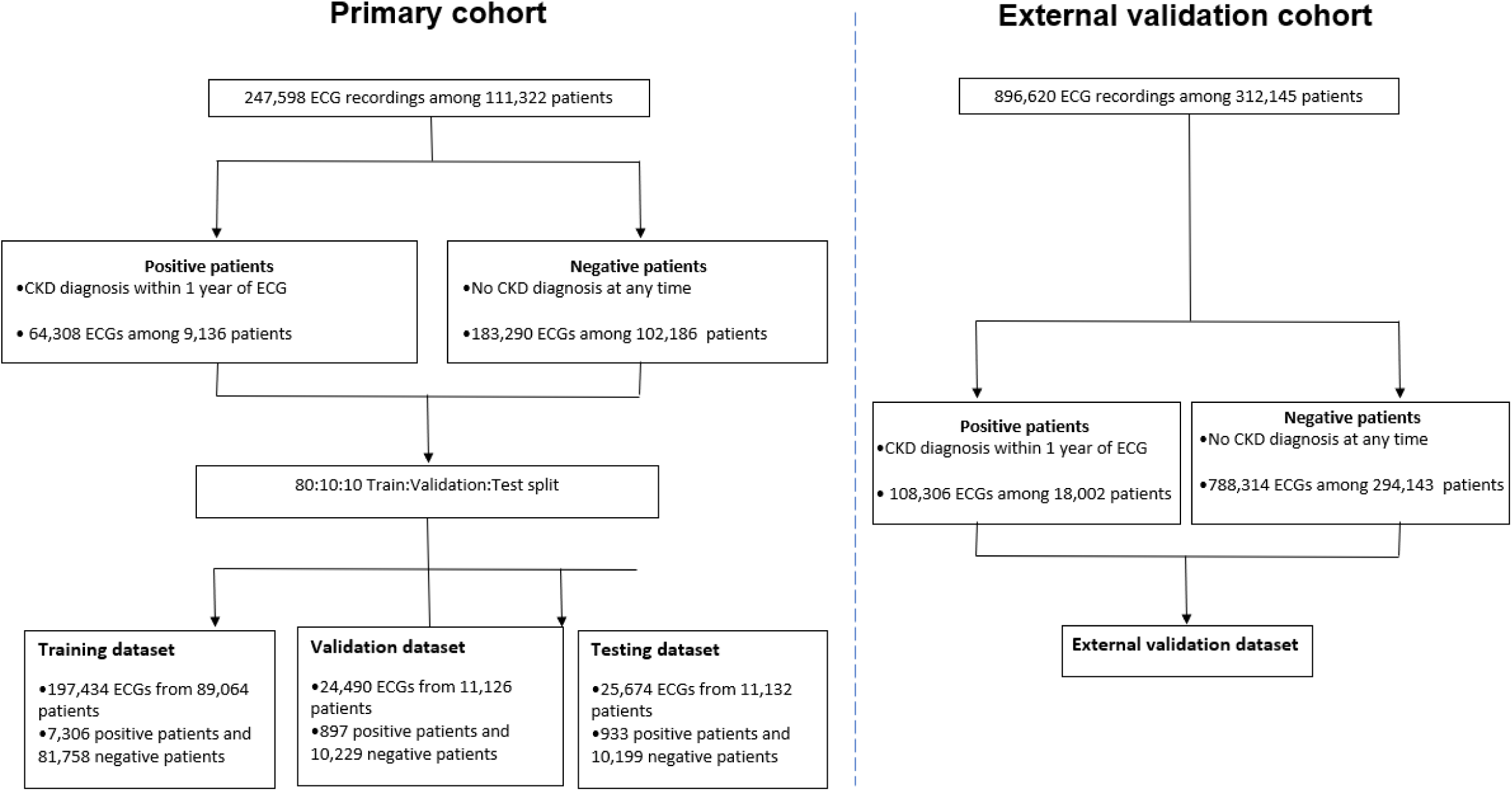
Study subject selection. CKD=Chronic kidney disease.

The ECG waveform data were acquired at a sampling rate of 500 Hz and extracted as 10 second, 12×5000 matrices of amplitude values. ECGs with missing leads were excluded from the study cohort. Associated clinical data for each patient was obtained from the electronic health record (EHR). Disease diagnoses were identified by International Classification of Diseases (ICD) 9^th^ edition codes and demographic and clinical characteristics (e.g., age, gender, BMI, cardiovascular disease) were also extracted from the electronic health records (Table 1). The institutional review boards of Cedars-Sinai Medical Center and Stanford Healthcare approved the study protocol.

**Table 1.** List of ICD9-codes that were used in the present study. CKD=Chronic kidney disease. ESRD=End-stage renal disease.

| **Disease** | **ICD9-code(s)** |
| --- | --- |
| Any stage CKD | 585.1+585.2+585.3+585.4+585.5+585.6 |
| Mild stage CKD | 585.1+585.2 |
| Moderate-severe CKD | 585.3+585.4+585.5 |
| ESRD | 585.6 |
| Hypertension | 401 |
| Diabetes Mellitus | 250 |
| Cardiovascular disease | 410-414+430-438 |
| Heart failure | 428 |
| Proteinuria | 791.0 |
| Anemia | 280-285 |

### AI Model Design and Training

We designed a novel convolutional neural network, for ECG interpretation with potential for clinical data integration to predict the primary outcomes of chronic kidney disease and end-stage renal disease (Figure 2). The model was trained to predict outcomes with the input of one 12-lead ECG obtained within 1 year of diagnosis. If the same patient had multiple ECGs, each was considered an independent case. Models were trained using the PyTorch deep learning framework. The model was initialized with random weights and trained using a binary cross-entropy loss function for up to 100 epochs with an ADAM optimizer and an initial learning rate of 1e-4. Early stopping was performed based on the validation dataset’s area under the receiver operating curve. Local Interpretable Model-agnostic Explanations (LIME)(15, 19) was used with 1000 samples per study to identify relevant features in the ECG waveform by iteratively randomly perturbing 0.5% of the waveform and identifying which changes most impacted model performance.

**Figure 2.**
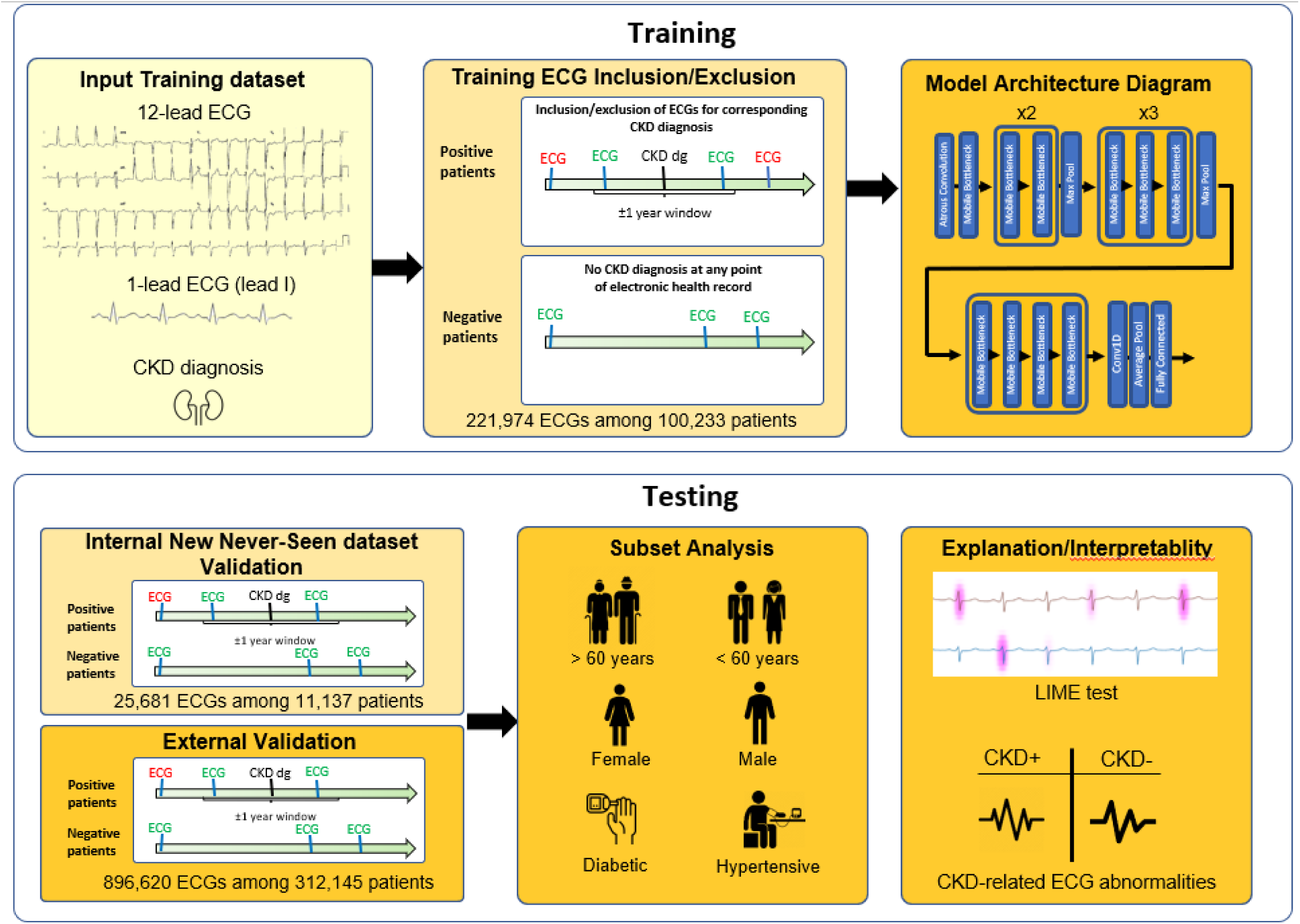
Schematic illustration of deep learning model training, testing, and validation. CKD=Chronic kidney disease.

### Statistical Analysis

All analyses were performed on the held-out test dataset, which was never seen during model training. The performance of the model in predicting the primary outcomes was mathematically assessed by the area under the curve (AUC) of the receiver operating characteristic (ROC) curve. After model derivation and training, primary and secondary analyses were performed on trained models using the held-out test cohort. Secondary sensitivity analyses were limited to procedures performed in patients with diabetes, hypertension, male, and age greater or lower than 60 years old. We computed two-sided 95% confidence intervals using 1,000 bootstrapped samples for each calculation. Statistical analysis was performed in R and Python.

## Results

### Primary cohort characteristics

Our primary cohort consisted of a total of 247,655 ECGs, of which 221,974 were randomized to the training set (for both training and validation) and 25,681 to the testing set. The number of patients in the training set was 100,233 of which 0.8% had mild CKD, 3.5% had moderate-severe CKD, and 2.7% had ESRD. The testing set included 11,137 patients of which 0.7% had mild CKD, 3.6% had moderate-severe CKD, and 2.8% had ESRD. The mean age of the primary cohort was 61.3 ±19.7 years and 48% were female. The proportion of Caucasians was 60.4%, whereas 13.8% were black, 5.5% were Asians, and 20.3% had other or unknown race. Demographic and clinical characteristics are presented in Table 2.

**Table 2.** Demographic and clinical characteristics in the internal and external dataset. Continuous variables are presented as mean±standard deviation. BMI=Body mass index, ESRD=End-stage renal disease.

| Characteristic | Internal training and validation datasets |  |  | External validation dataset |
| --- | --- | --- | --- | --- |
|  | Total | Training | Test |  |
| Number of patients | 111,370 | 100,233 | 11,137 | 312,145 |
| Number of ECGs | 247,655 | 221,974 | 25,681 | 896,620 |
| **<u>Demographics</u>** |  |  |  |  |
| Age, years | 61.3 ±19.7 | 61.3 ±19.6 | 61.5 ±19.9 | 56.7±18.7 |
| Female, n (%) | 53,476 (48.0%) | 48,133 (48.0%) | 5,343 (48.0%) | 139,235 (50.4%) |
| BMI, kg/m<sup>2</sup> | 26.7±16.6 | 26.7 ±18.2 | 26.6 ±7.2 | 27.6±6.6 |
| Caucasian, n (%) | 67,253 (60.4%) | 60,550 (60.4%) | 6,703 (60.2%) | 148,367 (47.5%) |
| Black, n (%) | 15,323 (13.8%) | 13,751 (13.7%) | 1,572 (14.1%) | 11,191 (3.6%) |
| Asian, n (%) | 6,135 (5.5%) | 5,558 (5.5%) | 577 (5.2%) | 38,229 (12.3%) |
| Other/unknown race, n (%) | 22,659 (20.3%) | 20,374 (20.3%) | 2,285 (20.5%) | 114,358 (36.6%) |
| **<u>Clinical characteristics, n (%)</u>** |  |  |  |  |
| Hypertension | 18,335 (16.5%) | 14,621 (16.4%) | 1,871 (16.8%) | 110,311 (35.3%) |
| Diabetes Mellitus | 8,728 (7.8%) | 6,970 (7.8%) | 914 (8.2%) | 14,152 (4.5%) |
| Cardiovascular disease | 15,719 (14.1%) | 12,561 (14.1%) | 1,577 (14.2%) | 34,582 (11.1%) |
| Heart Failure | 9,604 (8.6%) | 7,634 (8.6%) | 995 (8.9%) | 20,167 (6.5%) |
| Proteinuria | 1,007 (0.9%) | 795 (0.9%) | 110 (1.0%) | 2,086 (0.7%) |
| Anemia | 11,933 (10.7%) | 9,537 (10.7%) | 1,194 (10.7%) | 18,653 (6.0%) |
| **<u>Chronic kidney disease, n (%)</u>** |  |  |  |  |
| Mild (Stage 1 -2) | 864 (0.8%) | 782 (0.8%) | 82 (0.7%) | 3,727 (1.2%) |
| Moderate (Stage 3-5) | 3918 (3.5%) | 3515 (3.5%) | 403 (3.6%) | 11,335 (3.6%) |
| ESRD | 3034 (2.7%) | 2722 (2.7%) | 312 (2.8%) | 2,940 (0.9%) |

### Model performance in the primary cohort

Our 12-lead ECG-based model achieved discrimination of any stage CKD with an AUC of 0.767 (95% CI 0.76-0.773). The model performance was consistent across the range of CKD stage, with our model achieving an AUC of 0.753 (0.735-0.770) in discriminating mild CKD, AUC of 0.759 (0.750-0.767) in discriminating moderate-severe CKD, and AUC of 0.783 (0.730-0.752) in discriminating ESRD. In all cases, negative examples were defined as ECGs without CKD diagnoses. Sensitivity and specificity at detecting any stage CKD were 0.699 (0.699-0.699) and 0.698 (0.698-0.698), respectively.

Given the increased prevalence of wearable technologies, particularly devices that include single lead ECG information, we trained an additional deep learning model with information from only single lead ECG information to simulate the DLA’s performance with single-lead wearable information. With 1-lead ECG waveform data, DLA achieved an AUC of 0.744 (0.737-0.751) in detecting any stage CKD, with sensitivity and specificity of 0.723 (0.723-0.723) and 0.643 (0.643-0.643), respectively. In addition, 1-lead ECG-based DLA achieved an AUC of 0.746 (0.728-0.764) in detecting mild CKD, AUC of 0.735 (0.726-0.744) in detecting moderate-severe CKD, and AUC of 0.757 (0.748-0.767) in detecting ESRD.

Since early detection of CKD is crucial to prevent disease progression and complications in older age, we tested the performance of our model in younger patients (<60 years of age). 12-lead and 1-lead ECG-based DLAs were able to detect any stage CKD with AUCs of 0.843 (0.834-0.852) and 0.824 (0.814-0.833) among patients under 60 years of age, respectively. Sensitivity in detecting any stage CKD was 0.761 (0.761-0.761) with 12-lead ECG waveform and 0.812 (0.812-0.812) with 1-lead ECG waveform. Specificities were 0.787 (0.787-0.787) and 0.705 (0.705-0.705) for 12-lead and 1-lead ECG waveforms, respectively.

We also tested the performance of our model separately among diabetic, hypertensive, older patients, who are generally considered as high-risk subgroups. 12-lead based model detected CKD with an AUC of 0.747 (0.707-0.783) among diabetic patients, an AUC of 0.714 (0.701-0.726) among patients with hypertension, and an AUC of 0.706 (0.697-0.716) among patients greater than 60 years old. Similarly, 1-lead ECG-based DLA achieved discrimination of any stage CKD among diabetic, hypertensive, and patients > 60 years with AUCs of 0.66<u>3</u> (0.625-0.707), 0.678 (0.663-0.691), and 0.681 (0.671-0.691), respectively. In addition, 12-lead and 1-lead ECG-based models detected CKD with similar accuracy among male (AUCs of 0.764 (0.755-0.772) and 0.742 (0.733-0.750), respectively) and female patients (AUCs of 0.756 (0.745-0.768) and 0.735 (0.723-0.747), respectively). Detailed results for 1-lead and 12-lead ECG-based DLA performance in the held-out test set are presented in Table 3 and Table 4, while AUC curves are illustrated in Figure 3.

**Table 3.** Performance of the 12-lead ECG-based deep learning algorithm in the internal dataset. AUC=area under the receiver operating characteristics curve, CI=Confidence interval, CKD=Chronic kidney disease, ESRD=End-stage renal disease. PPV=positive predictive value. NPV=negative predictive value.

| Model | AUC<br>(95% CI) | Sensitivity<br>(95% CI) | Specificity<br>(95% CI) | PPV<br>(95% CI) | NPV<br>(95% CI) |
| --- | --- | --- | --- | --- | --- |
| **12-lead ECG models** |  |  |  |  |  |
| Any stage CKD | 0.767 (0.76-0.773) | 0.699 (0.699-0.699) | 0.698 (0.698-0.698) | 0.443 (0.443-0.443) | 0.871 (0.871-0.871) |
| Mild CKD | 0.753 (0.735-0.77) | 0.75 (0.75-0.75) | 0.644 (0.644-0.644) | 0.064 (0.064-0.064) | 0.987 (0.987-0.987) |
| Moderate-severe CKD | 0.759 (0.75-0.767) | 0.785 (0.785-0.785) | 0.598 (0.598-0.598) | 0.271 (0.271-0.271) | 0.936 (0.936-0.936) |
| ESRD | 0.783 (0.773-0.793) | 0.704 (0.704-0.704) | 0.726 (0.726-0.726) | 0.237 (0.237-0.237) | 0.953 (0.953-0.953) |
| **High Risk Subgroup analyses for any stage CKD** |  |  |  |  |  |
| Diabetic patients | 0.747 (0.707-0.783) | 0.699 (0.699-0.699) | 0.682 (0.682-0.682) | 0.906 (0.906-0.906) | 0.342 (0.342-0.342) |
| Hypertensive patients | 0.714 (0.701-0.726) | 0.659 (0.659-0.659) | 0.66 (0.66-0.66) | 0.798 (0.798-0.798) | 0.487 (0.487-0.487) |
| Age > 60 years | 0.706 (0.697-0.716) | 0.604 (0.604-0.604) | 0.701 (0.701-0.701) | 0.397 (0.397-0.397) | 0.844 (0.844-0.844) |
| Male | 0.764 (0.755-0.772) | 0.727 (0.727-0.727) | 0.666 (0.666-0.666) | 0.485 (0.485-0.485) | 0.849 (0.849-0.849) |
| Female | 0.756 (0.745-0.768) | 0.699 (0.699-0.699) | 0.680 (0.680-0.680) | 0.351 (0.351-0.351) | 0.902 (0.902-0.902) |
| **Screening cohort (age < 60)** |  |  |  |  |  |
| Any stage CKD | 0.843 (0.836-0.852) | 0.761 (0.761-0.761) | 0.787 (0.787-0.787) | 0.57 (0.57-0.57) | 0.899 (0.899-0.899) |
| Mild CKD | 0.795 (0.766-0.823) | 0.702 (0.702-0.702) | 0.748 (0.748-0.748) | 0.075 (0.075-0.075) | 0.989 (0.989-0.989) |
| Moderate-severe CKD | 0.854 (0.842-0.865) | 0.792 (0.792-0.792) | 0.787 (0.787-0.787) | 0.373 (0.373-0.373) | 0.959 (0.959-0.959) |
| ESRD | 0.842 (0.831-0.853) | 0.746 (0.746-0.746) | 0.791 (0.791-0.791) | 0.394 (0.394-0.394) | 0.945 (0.945-0.945) |

**Table 4.** Performance of the 1-lead ECG-based deep learning algorithm in the internal dataset. AUC=area under the receiver operating characteristics curve, CI=Confidence interval, CKD=Chronic kidney disease, ESRD=End-stage renal disease. PPV=positive predictive value. NPV=negative predictive value.

| Model | AUC<br>(95% CI) | Sensitivity<br>(95% CI) | Specificity<br>(95% CI) | PPV<br>(95% CI) | NPV<br>(95% CI) |
| --- | --- | --- | --- | --- | --- |
| **1-lead ECG models** |  |  |  |  |  |
| Any stage CKD | 0.744 (0.737-0.751) | 0.723 (0.723-0.723) | 0.643 (0.643-0.643) | 0.41 (0.41-0.41) | 0.871 (0.871-0.871) |
| Mild CKD | 0.746 (0.728-0.764) | 0.735 (0.735-0.735) | 0.66 (0.66-0.66) | 0.066 (0.066-0.066) | 0.987 (0.987-0.987) |
| Moderate-severe CKD | 0.735 (0.726-0.744) | 0.732 (0.732-0.732) | 0.618 (0.618-0.618) | 0.267 (0.267-0.267) | 0.924 (0.924-0.924) |
| ESRD | 0.757 (0.748-0.767) | 0.738 (0.738-0.738) | 0.647 (0.647-0.647) | 0.202 (0.202-0.202) | 0.953 (0.953-0.953) |
| **High Risk Subgroup analyses for any stage CKD** |  |  |  |  |  |
| Diabetic patients | 0.663 (0.625-0.707) | 0.819 (0.819-0.819) | 0.457 (0.457-0.457) | 0.868 (0.868-0.868) | 0.367 (0.367-0.367) |
| Hypertensive patients | 0.678 (0.663-0.691) | 0.713 (0.713-0.713) | 0.555 (0.555-0.555) | 0.765 (0.765-0.765) | 0.487 (0.487-0.487) |
| Age > 60 years | 0.681 (0.671-0.691) | 0.679 (0.679-0.679) | 0.588 (0.588-0.588) | 0.35 (0.35-0.35) | 0.849 (0.849-0.849) |
| Male | 0.742 (0.733-0.75) | 0.747 (0.747-0.747) | 0.612 (0.612-0.612) | 0.455 (0.455-0.455) | 0.848 (0.848-0.848) |
| Female | 0.735 (0.723-0.747) | 0.672 (0.672-0.672) | 0.681 (0.681-0.681) | 0.342 (0.342-0.342) | 0.894 (0.894-0.894) |
| **Screening cohort (age < 60)** |  |  |  |  |  |
| Any stage CKD | 0.824 (0.815-0.832) | 0.812 (0.812-0.812) | 0.705 (0.705-0.705) | 0.506 (0.506-0.506) | 0.91 (0.91-0.91) |
| Mild CKD | 0.819 (0.791-0.844) | 0.771 (0.771-0.771) | 0.758 (0.758-0.758) | 0.085 (0.085-0.085) | 0.991 (0.991-0.991) |
| Moderate-severe CKD | 0.828 (0.816-0.84) | 0.814 (0.814-0.814) | 0.713 (0.713-0.713) | 0.313 (0.313-0.313) | 0.96 (0.96-0.96) |
| ESRD | 0.82 (0.808-0.831) | 0.82 (0.82-0.82) | 0.693 (0.693-0.693) | 0.328 (0.328-0.328) | 0.955 (0.955-0.955) |

**Figure 3.**
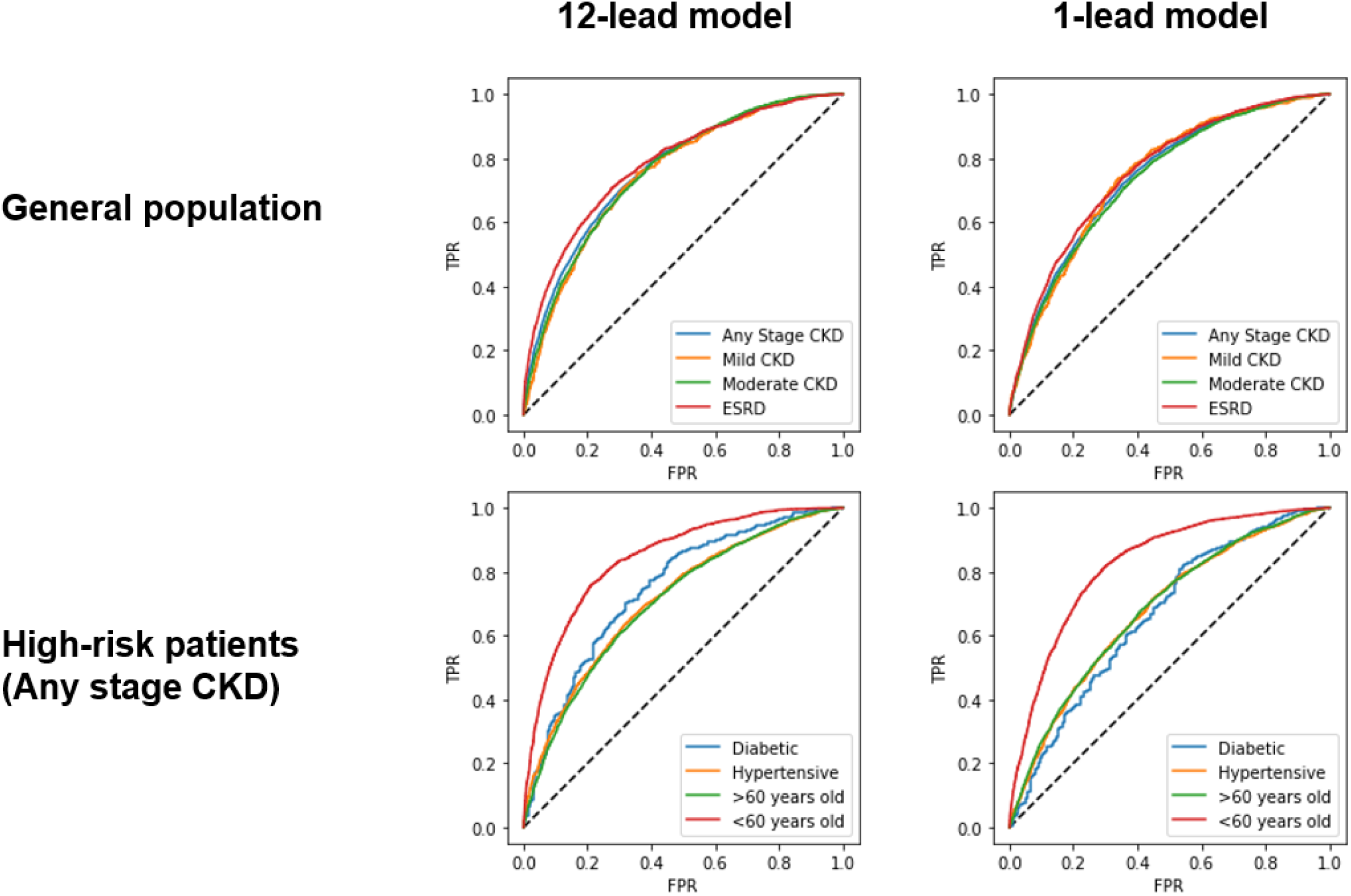
Model performance in the internal dataset. CKD=Chronic kidney disease. ESRD=End-stage renal disease.

### Electrocardiographic features in CKD

To understand the key features of relevance for our deep learning model to be able to detect CKD, we performed two sets of experiments to evaluate the ECG parameters that are important for identifying CKD. We found statistically significant differences in all available ECG variables (heart rate, PR interval, P wave duration, QRS duration, QTc interval, P-wave axis, R-wave axis, T-wave axis) between CKD stages (Table 5). Most prominently, patients with CKD had prolonged PR interval, prolonged QRS duration, prolonged QTc interval, and skewed T wave axis in comparison to those without CKD.

**Table 5.**
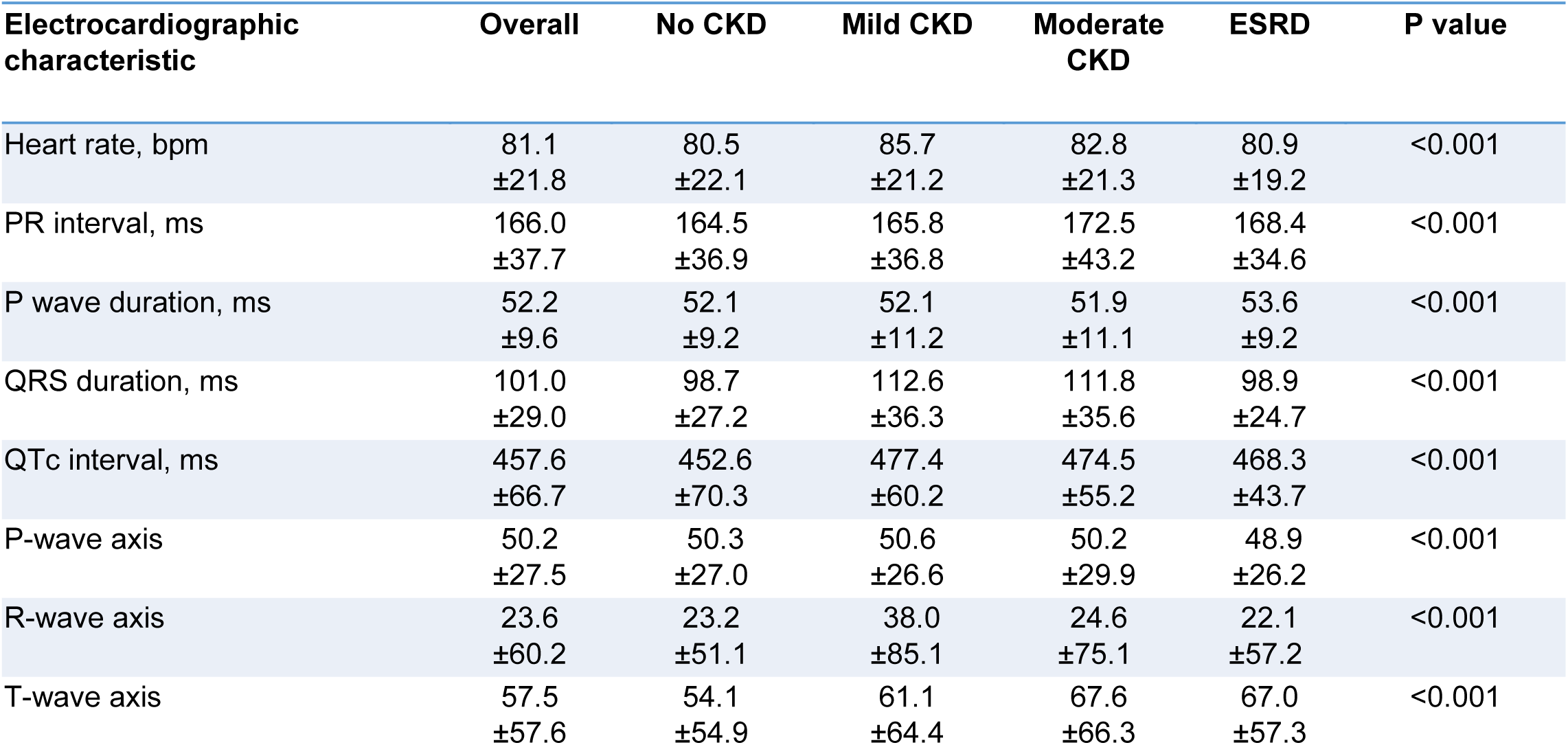
Electrocardiographic characteristics according to chronic kidney disease (CKD) stage in the training dataset. Data is presented as mean±standard deviation. ESRD=End-stage renal disease.

| Electrocardiographic characteristic | Overall | No CKD | Mild CKD | Moderate CKD | ESRD | P value |
| --- | --- | --- | --- | --- | --- | --- |
| Heart rate, bpm | 81.1<br>±21.8 | 80.5<br>±22.1 | 85.7<br>±21.2 | 82.8<br>±21.3 | 80.9<br>±19.2 | <0.001 |
| PR interval, ms | 166.0<br>±37.7 | 164.5<br>±36.9 | 165.8<br>±36.8 | 172.5<br>±43.2 | 168.4<br>±34.6 | <0.001 |
| P wave duration, ms | 52.2<br>±9.6 | 52.1<br>±9.2 | 52.1<br>±11.2 | 51.9<br>±11.1 | 53.6<br>±9.2 | <0.001 |
| QRS duration, ms | 101.0<br>±29.0 | 98.7<br>±27.2 | 112.6<br>±36.3 | 111.8<br>±35.6 | 98.9<br>±24.7 | <0.001 |
| QTc interval, ms | 457.6<br>±66.7 | 452.6<br>±70.3 | 477.4<br>±60.2 | 474.5<br>±55.2 | 468.3<br>±43.7 | <0.001 |
| P-wave axis | 50.2<br>±27.5 | 50.3<br>±27.0 | 50.6<br>±26.6 | 50.2<br>±29.9 | 48.9<br>±26.2 | <0.001 |
| R-wave axis | 23.6<br>±60.2 | 23.2<br>±51.1 | 38.0<br>±85.1 | 24.6<br>±75.1 | 22.1<br>±57.2 | <0.001 |
| T-wave axis | 57.5<br>±57.6 | 54.1<br>±54.9 | 61.1<br>±64.4 | 67.6<br>±66.3 | 67.0<br>±57.3 | <0.001 |

Secondly, we used LIME to identify which ECG segments were particularly used in the identification of CKD. Figure 4 shows examples of LIME-highlighted ECG segments in 12-lead and 1-lead ECG waveforms taken from correctly recognized CKD and healthy control patients in the held-out test set. In both examples, the LIME-highlighted ECG features focused mostly on QRS complexes and PR intervals. In addition, QRS complexes and PR intervals in limb leads were most frequently highlighted, potentially denoting CKD-associated electrophysiologic alterations.

**Figure 4.**
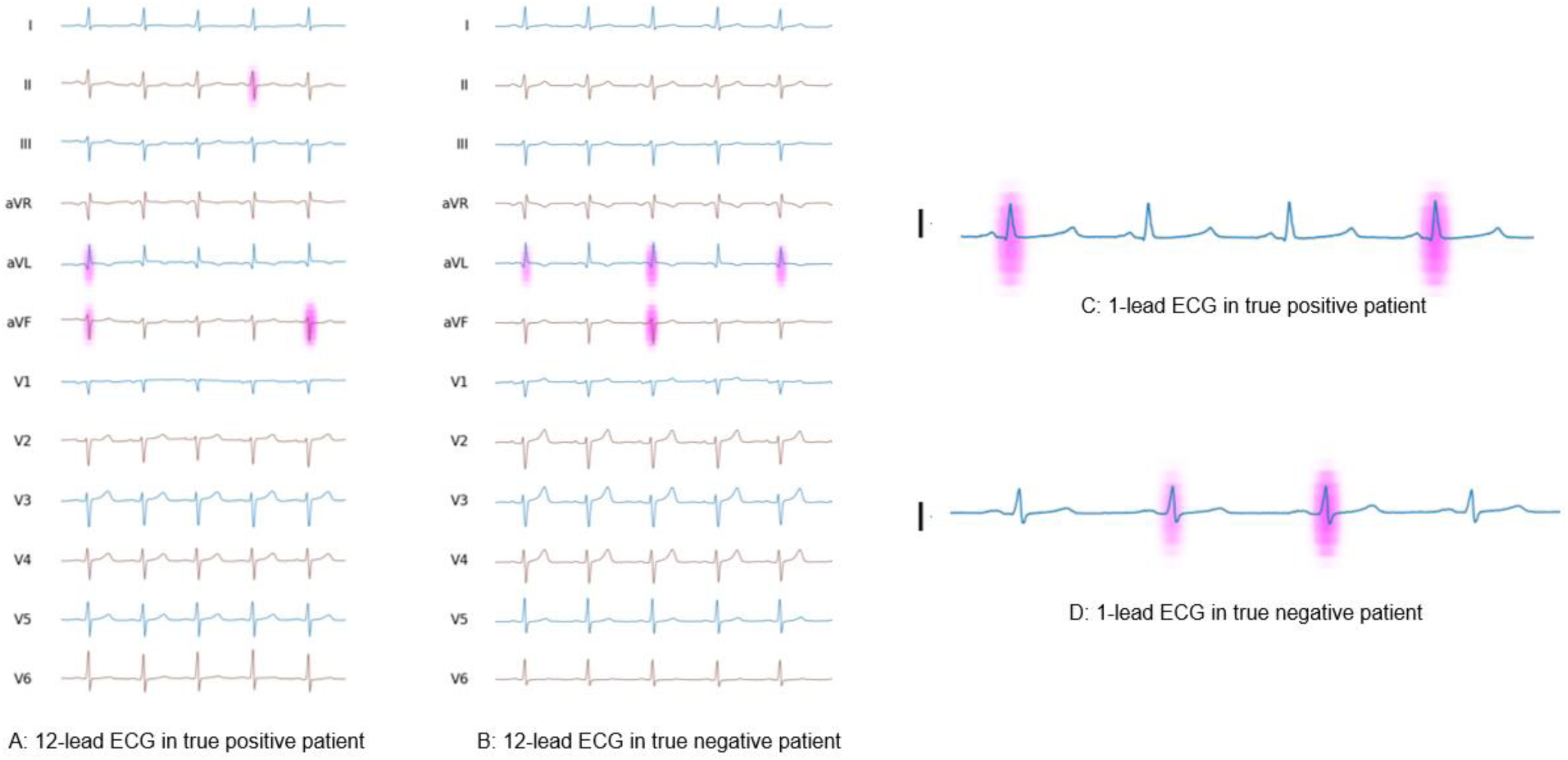
The Linear Interpretable Model-Agnostic Explanations (LIME) map of 12-lead and 1-lead ECGs highlights features that were important for deep learning model in chronic kidney disease detection. Important ECG features are highlighted with greater color intensity. In both true positive (A and C) and true negative patients (B and D) LIME highlighted QRS complexes and PR intervals.

### External validation cohort characteristics

The external validation cohort consisted of a total of 896,620 ECGs among 312,145 patients. The prevalence of mild CKD was 1.2% while 3.6% had moderate-severe CKD, and 0.9% had ESRD. The mean age of the external validation cohort was 56.7±18.7 years and 50.4% were female. The proportion of Caucasians was 47.5%, while 3.6% were black, 12.3% were Asians, and 36.6% had other or unknown race. Demographic and clinical characteristics are presented in Table 1.

### Model performance in the external validation dataset

In the external validation dataset, our 12-lead and 1-lead models’ performances were comparable to the primary cohort. 12-lead ECG-based model achieved an AUC of 0.709 (0.708-0.710) in discriminating any stage CKD, with a sensitivity of 0.212 (0.210-0.214) and specificity of 0.926 (0.926-0.927). Additionally, 12-lead ECG-based model achieved an AUC of 0.679 (0.675-0.682) in discriminating mild CKD, AUC of 0.714 (0.713-0.716) in discriminating moderate-severe CKD, and AUC of 0.767 (0.764-0.769) in discriminating ESRD. 1-lead ECG-based model detected any stage CKD with an AUC of 0.701 (0.700-0.702), mild stage CKD with an AUC of 0.671 (0.668-0.674), moderate-severe CKD with an AUC of 0.694 (0.692-0.695), and ESRD with an AUC of 0.780 (0.778-0.782).

Consistent with the primary cohort in which our model achieved higher CKD detection accuracy among younger patients, 12-lead and 1-lead ECG-based models achieved AUCs of 0.784 (0.782-0.786) and 0.777 (0.775-0.779) in detecting any stage CKD among subjects under 60 years of age, respectively. However, the 12-lead ECG-based model’s CKD discrimination accuracy was somewhat lower in high-risk subgroups with an AUC of 0.699 (0.697-0.702) among diabetic, AUC of 0.712 (0.710-0.714) among hypertensive, and AUC of 0.660 (0.658-0.661) among subjects over 60 years of age. Detailed results for 1-lead and 12-lead ECG-based DLA performance in the external validation cohort are presented in Table 6 and Table 7.

**Table 6.** Performance of the 12-lead ECG-based deep learning algorithm in the external dataset. AUC=area under the receiver operating characteristics curve, CI=Confidence interval, CKD=Chronic kidney disease, ESRD=End-stage renal disease. PPV=positive predictive value. NPV=negative predictive value.

| Model | AUC<br>(95% CI) | Sensitivity<br>(95% CI) | Specificity<br>(95% CI) | PPV<br>(95% CI) | NPV<br>(95% CI) |
| --- | --- | --- | --- | --- | --- |
| **12-lead ECG models** |  |  |  |  |  |
| Any stage CKD | 0.709 (0.708-0.710) | 0.573 (0.571-0.575) | 0.720 (0.719-0.721) | 0.276 (0.275-0.278) | 0.900 (0.900-0.901) |
| Mild CKD | 0.679 (0.675-0.682) | 0.525 (0.519-0.530) | 0.720 (0.719-0.721) | 0.047 (0.046-0.048) | 0.983 (0.983-0.983) |
| Moderate-severe CKD | 0.714 (0.713-0.716) | 0.585 (0.582-0.588) | 0.720 (0.719-0.720) | 0.141 (0.140-0.143) | 0.956 (0.956-0.957) |
| ESRD | 0.767 (0.764-0.769) | 0.665 (0.660-0.670) | 0.720 (0.719-0.721) | 0.083 (0.082-0.084) | 0.983 (0.982-0.983) |
| **High Risk Subgroup analyses for any stage CKD** |  |  |  |  |  |
| Diabetic patients | 0.699 (0.697-0.702) | 0.620 (0.615-0.625) | 0.670 (0.665-0.674) | 0.711 (0.708-0.716) | 0.573 (0.568-0.577) |
| Hypertensive patients | 0.712 (0.710-0.714) | 0.555 (0.552-0.558) | 0.741 (0.740-0.742) | 0.378 (0.375-0.380) | 0.855 (0.854-0.856) |
| Age > 60 years | 0.660 (0.658-0.661) | 0.582 (0.579-0.584) | 0.648 (0.647-0.649) | 0.349 (0.347-0.350) | 0.827 (0.826-0.828) |
| Male | 0.719 (0.718-0.721) | 0.593 (0.590-0.596) | 0.721 (0.720-0.723) | 0.387 (0.384-0.389) | 0.857 (0.856-0.858) |
| Female | 0.728 (0.726-0.730) | 0.537 (0.533-0.541) | 0.769 (0.768-0.770) | 0.279 (0.276-0.281) | 0.909 (0.908-0.910) |
| **Screening cohort (age < 60)** |  |  |  |  |  |
| Any stage CKD | 0.784 (0.782-0.786) | 0.552 (0.548-0.556) | 0.843 (0.843-0.845) | 0.326 (0.323-0.328) | 0.932 (0.931-0.933) |
| Mild CKD | 0.763 (0.757-0.767) | 0.524 (0.516-0.536) | 0.843 (0.843-0.844) | 0.063 (0.061-0.065) | 0.989 (0.989-0.989) |
| Moderate-severe CKD | 0.780 (0.777-0.782) | 0.548 (0.543-0.556) | 0.843 (0.843-0.844) | 0.128 (0.126-0.130) | 0.978 (0.978-0.979) |
| ESRD | 0.841 (0.839-0.844) | 0.649 (0.644-0.656) | 0.843 (0.843-0.845) | 0.155 (0.153-0.158) | 0.982 (0.982-0.982) |

**Table 7.**
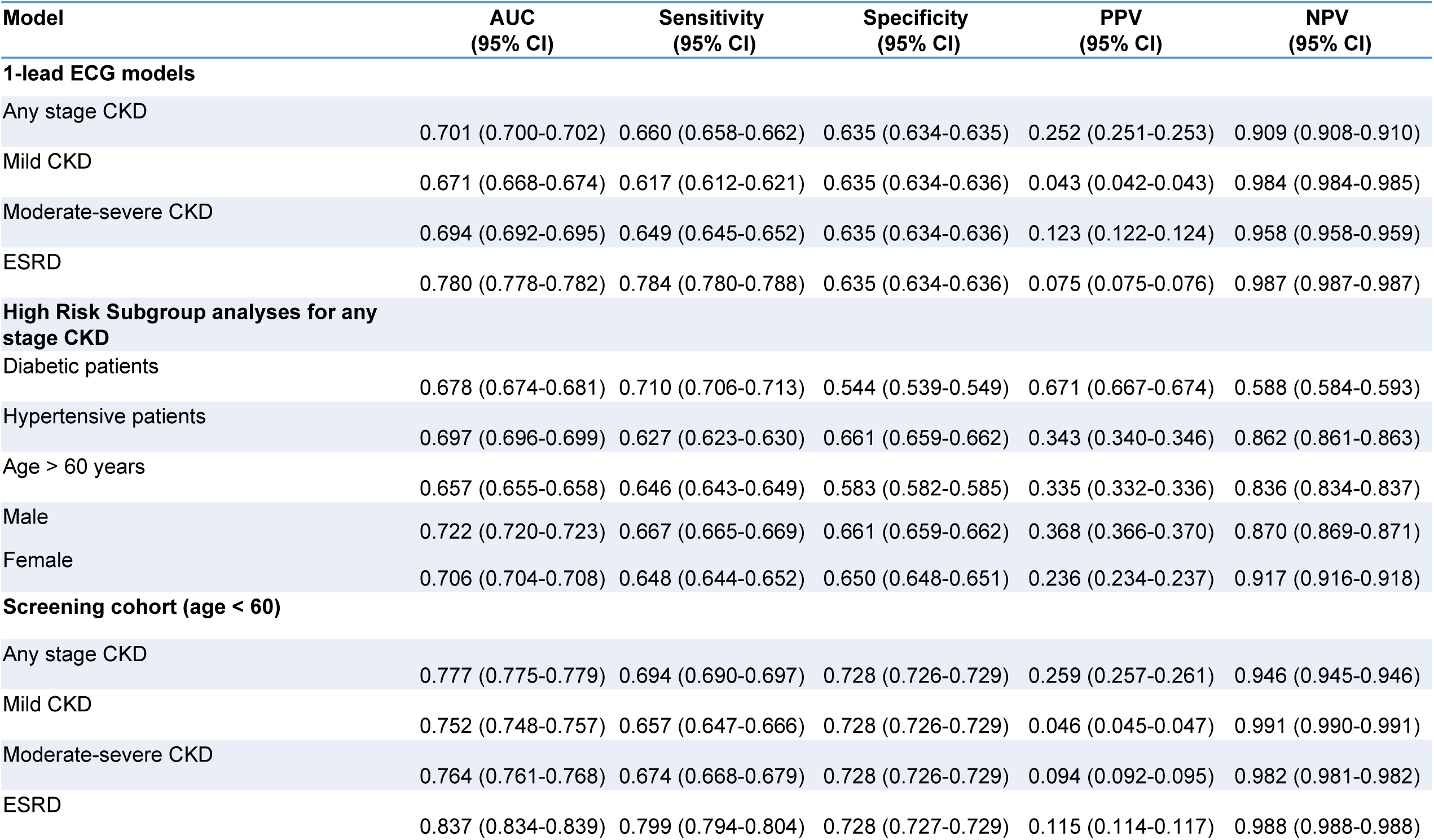
Performance of the 1-lead ECG-based deep learning algorithm in the external dataset. AUC=area under the receiver operating characteristics curve, CI=Confidence interval, CKD=Chronic kidney disease, ESRD=End-stage renal disease. PPV=positive predictive value. NPV=negative predictive value.

## Discussion

In the present study, we investigated the performance of a novel deep learning model to detect CKD using ECG waveforms. Our 12-lead ECG-based model had good accuracy in identifying any stage CKD and higher accuracy in detecting CKD in patients under 60 years of age. Accuracy also improved along with the worsening CKD stage. These results were validated in a separate health care system, that also showed good discrimination accuracy for the presence of any stage CKD in the whole study population and higher discrimination accuracy among patients under 60 years of age. While 12-lead ECGs are widely available in the healthcare unit settings, rapid adoption of wearable technology has also introduced opportunities for large-scale data collection outside of formal healthcare settings. Our 1-lead ECG-based DLA showed good discrimination accuracy for CKD in young patients, suggesting artificial intelligence may possess significant potential in widescale screening in this patient population. One-lead ECGs could also increase screening rates in high-risk patients (Figure 5 and Figure 6).

**Figure 5.**
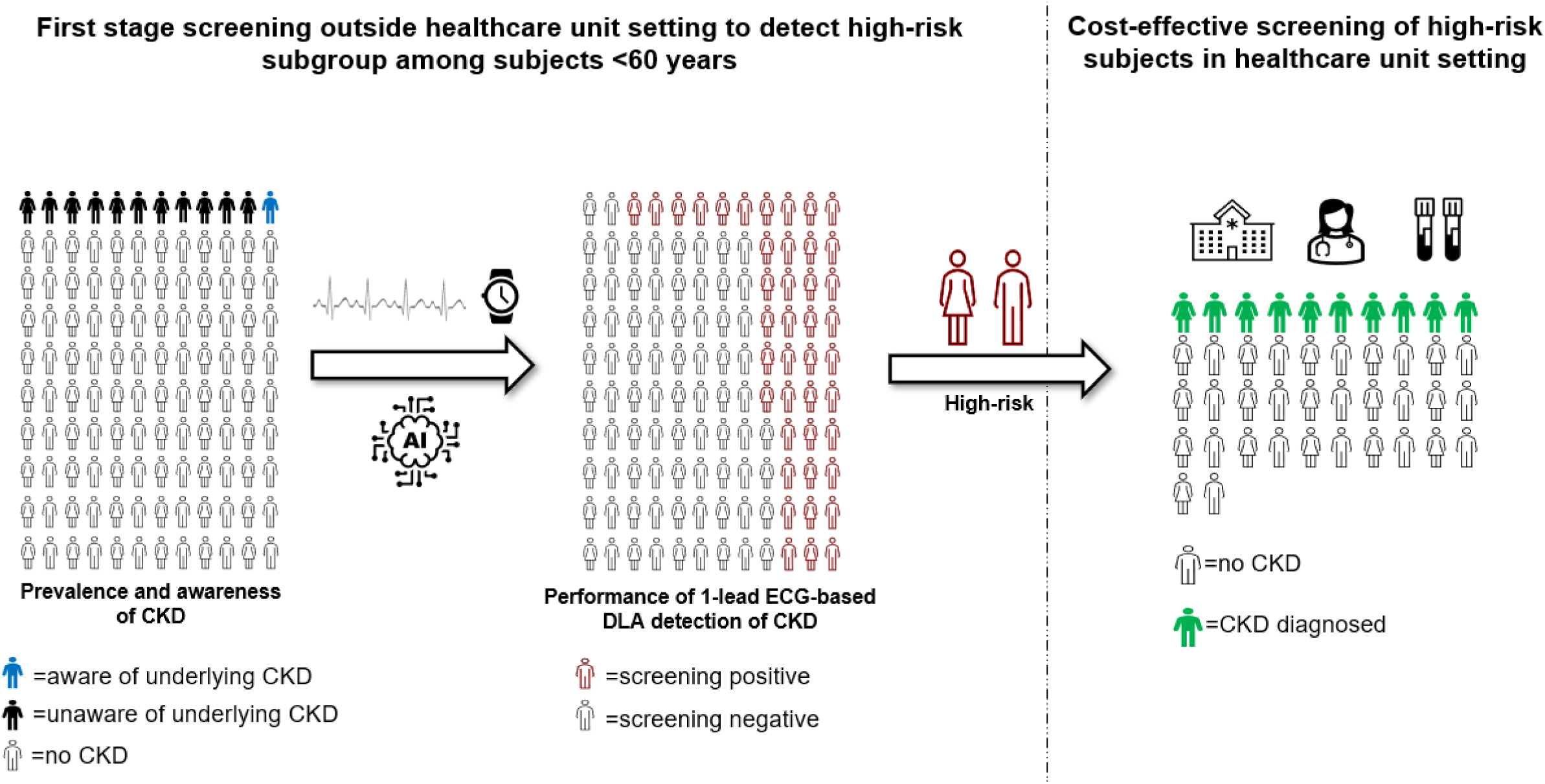
Schematic illustration of utilizing ECG-based deep learning model to detect a high-risk subgroup of subjects under 60 years of age for chronic kidney disease (CKD) screening. DLA=Deep learning algorithm.

**Figure 6.**
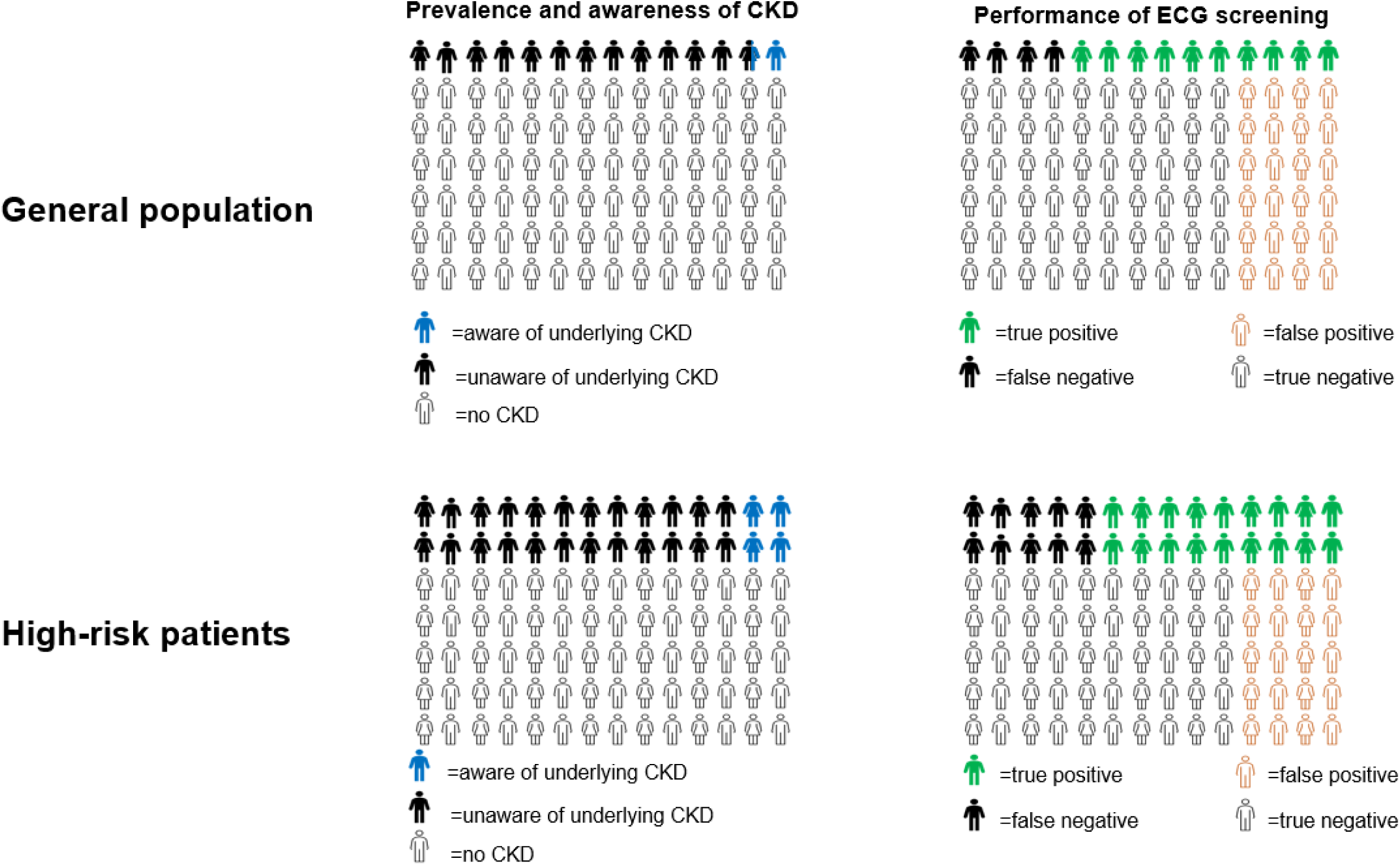
Illustration of chronic kidney disease (CKD) prevalence and awareness in comparison to model performance.

Low awareness of CKD and limitations in current screening measures highlight the urgency of novel screening strategies to increase detection rates of early-stage CKD. Previous studies have demonstrated that the cost-effectiveness of CKD screening is highly dependent on patient risk factor profile and CKD probability, and there has been debate on whether CKD screening should be targeted only to high-risk patients, or also extend to patients without risk factors for CKD(20-23). Although screening high-risk patients is guideline-recommended, testing rates remain low as only about 20% of high-risk patients receive guideline-recommended assessment in the U.S.(24). Consequently, most of the high-risk patients are likely to be unaware of underlying CKD(2, 3).

Our model performed better at detecting CKD in younger patients, whereas detection accuracy was lower in older and high-risk patients. Reasons for this observation are not fully clear but may be due to the fact that younger patients in general have fewer comorbidities, meaning that any detected ECG abnormalities may be especially meanintgul and specific.. Although older age is a well-known risk marker for CKD, the prevalence of CKD in younger patients is also notably high in the U.S. (8-10% in <65 years)(3). Remarkably, however, awareness of underlying CKD is also very low in younger patients, as only about 8% are aware of the disease(3). Given the availability of effective low-risk CKD treatments and the reversibility of CKD, there are substantial potential benefits for detecting and treating CKD, especially in the young.

The strengths of our study include the large cohort of patients undergoing ECG recording across a decade and the use of state-of-the-art deep learning architectures. We also used two separate approaches to understand the key features of relevance for our deep learning model. While previous studies have reported that patients with CKD have high rates of P wave abnormalities, prolonged PR interval, QTc prolongation, QT dispersion, and LVH(25-28), in the present study CKD was associated with skewed P-, R-, and T-wave axes in addition to prolonged QRS, PR, and QTc intervals. However, a few limitations warrant consideration. Our study is retrospective, and study populations are derived from two large academic medical centers situated in dense urban metropolitan areas. Validation in prospective general population cohorts in outpatient settings is required to confirm an ECG-based DLA’s ability to recognize patients with CKD. Moreover, the prevalence of mild CKD was low in our cohort and we cannot exclude the possibility that some of the study subjects without CKD diagnosis in electronic health records have an undiagnosed disease, as especially mild-stage CKD can often be undiagnosed, particularly using an ICD9 code-based adjudication.

By 2030, the UN’s Sustainable Development Goals are to reduce premature mortality related to non-communicable diseases by a third. Given the high prevalence of asymptomatic CKD, serious consequences of untreated disease, presence of effective low-risk treatment, and detectable preclinical state with inexpensive and simple diagnostic tests, CKD represents a good target for large-scale population screening and harbors the potential for reducing premature mortality related to non-communicable diseases. In addition to the high mortality and morbidity due to CKD, treatment costs for CKD are also high and have increased during the last decades(29). Especially, the increasing number of patients requiring renal replacement creates challenges for health care systems worldwide, and the shortage of sufficient replacement services may cause at least 2 million premature deaths annually(30). Therefore, widely available, inexpensive, and effective CKD prevention and management strategies are warranted to enable equal opportunities in reducing CKD-related disability-adjusted life years.

## Conclusions

Our ECG-based deep learning model was able to detect CKD with good discrimination accuracy in multiple study populations and with particularly high accuracy in patients under 60 years of age. These results suggest that deep learning-based ECG analysis may provide additional value in detecting various CKD stages, especially in younger patients. The clinical significance of this study lies in the importance of novel screening methods for early detection of CKD, which is crucial to enable early treatment and prevent disease progression.

## Data Availability

All data produced in the present study are available upon reasonable request to the authors.

## Contributors

Study design and conception: LH, MC, SSC, and DO. Acquisition, analysis or interpretation of data: LH, MC, NY, JWH, JT, MJ, PF, AK, RKS, JE, SC, JZ, SSC, and DO. Drafting of the manuscript: LH, MC, and DO. Statistical analysis: MC, JWH, and DO. Critical revision of the manuscript for important intellectual content: NY, JWH, JT, MJ, PF, AK, RKS, JE, SC, JZ, and SSC. Administrative and material support: SSC and DO. Obtained funding: DO. Supervision: SSC and DO. Full access to the data: MC and DO. All authors reviewed and approved the final version of the manuscript.

